# Using contact network dynamics to implement efficient interventions against pathogen spread in hospital settings

**DOI:** 10.1101/2023.12.08.23299666

**Authors:** Quentin Leclerc, Audrey Duval, Didier Guillemot, Lulla Opatowski, Laura Temime

**Affiliations:** Institut Pasteur, Université Paris Cité, Epidemiology and Modelling of Bacterial Escape to Antimicrobials (EMEA), 75015 Paris, France; INSERM, Université Paris-Saclay, Université de Versailles St-Quentin-en-Yvelines, Team Echappement aux Anti-infectieux et Pharmacoépidémiologie U1018, CESP, 78000 Versailles, France; Laboratoire Modélisation, Epidémiologie et Surveillance des Risques Sanitaires, Conservatoire National des Arts et Métiers, 75003 Paris, France; AP-HP, Paris Saclay, Department of Public Health, Medical Information, Clinical research, F-92380, Garches; Institut Pasteur, Conservatoire National des Arts et Métiers, Unité PACRI, 75015 Paris, France

**Keywords:** long-term care facility, contact network, individual-based model, nosocomial pathogens, interventions

## Abstract

Long-term care facilities (LTCF) are hotspots for pathogen transmission. Infection control interventions are essential, but the high density and heterogeneity of inter-individual contacts within LTCF may hinder their efficacy. Here, we explore how the patient-staff contact structure may inform effective intervention implementation. Using an individual-based model, we reproduced methicillin-resistant *Staphylococcus aureus* colonisation dynamics over a detailed contact network recorded within an LTCF, and examined the potential impact of three types of interventions against transmission (reallocation reducing the number of unique contacts per staff, reinforced contact precautions, and vaccination protecting against acquisition), targeted towards specific populations. All three interventions were effective when applied to all nurses or healthcare assistants (median reduction in MRSA colonisation incidence up to 21%), but the benefit did not exceed 8% when targeting any other single staff category. We identified “supercontactor” individuals with most contacts (“frequency-based”, overrepresented amongst nurses, porters and rehabilitation staff) or with the longest cumulative time spent in contact (“duration-based”, overrepresented amongst healthcare assistants and geriatric and persistent-vegetative-state patients). Targeting supercontactors enhanced interventions against pathogen spread in the LTCF. With contact precautions, targeting frequency-based staff supercontactors led to the highest incidence reduction (13%). Vaccinating duration-based patient supercontactors led to a higher reduction (22%) than all other approaches. Targeting supercontactors remained the most effective strategy when varying epidemiological parameters, indicating this approach can be broadly applied to prevent transmission of other nosocomial pathogens. Importantly, both staff and patients may be supercontactors, highlighting the importance of including patients in measures to prevent pathogen transmission in LTCF.

**Classification:** Biological Sciences, Biophysics and Computational Biology

**Significance statement:** Pathogen transmission is a challenge in long-term care facilities (LTCF) due to frequent and heterogeneous contacts of staff and patients. By characterising this contact structure and understanding the categories of staff and patients more likely to be “supercontactors”, with either more or longer contacts than others, we can implement more efficient interventions against pathogen spread. We illustrate this using a mathematical model to reproduce transmission of methicillin-resistant *Staphylococcus aureus* across a detailed contact network recorded in a LTCF. We show how the most efficient implementation strategy depends on the intervention (reallocation, contact precautions, vaccination) and target population (staff, patients, supercontactors). By varying epidemiological parameters, we demonstrate that these results are broadly applicable to prevent transmission of other nosocomial pathogens.

## Introduction

Healthcare associated infections (HAI) are a major threat worldwide, with more than 4 million infections occurring each year in Europe [1]. The recent COVID19 pandemic has underlined the high risk of pathogen dissemination in health care settings, similarly to what was previously reported for other coronaviruses, seasonal influenza or Ebola epidemics [2,3]. Bacterial nosocomial outbreaks are also frequently described, becoming more and more difficult to control with the rise of multidrug resistance [4]. In addition to significantly impacting the morbidity and mortality of hospitalized patients and potentially healthcare workers (HCWs), HAI generate additional costs due to longer hospital stays or additional expensive therapeutics, as well as legal consequences for practitioners and healthcare settings in case of patient lawsuits.

Methicillin-resistant *Staphylococcus aureus* (MRSA) is an important cause of such HAI, as these infections most often affect individuals in a weakened immunological state, such as hospitalized patients [5]. Crucially, MRSA colonization is a risk factor for infection, since individuals are more likely to be infected by a *S. aureus* strain they are carrying [6]. Consequently, it is essential to understand how individuals become colonized by MRSA in healthcare settings and to control the acquisition risk.

To limit pathogen dissemination through human cross-transmission in healthcare settings, a range of measures can be implemented, mostly based on improving contact precautions, such as patient isolation, hand-washing, wearing of gloves or masks. Vaccines to reduce the risk of pathogen colonisation also represent ongoing research and development topics, although none are commercially available and there have only been limited attempts to evaluate their impact in healthcare settings thus far [7]. However, the high density of human contacts involving HCWs, patients, and visitors, combined with variations in individual behaviours and overall stochasticity in transmission often limit the impact of these control measures. For instance, while efficient in general, hand-washing may fail due to a few “super-spreader” individuals who do not comply with hygiene recommendations [8].

Because the structure of contact networks within healthcare settings influences the spread of HAI pathogens [9], manipulating contact network structures or targeting highly connected individuals may significantly improve the efficacy of control measures [10]. Here, using individual-based modelling of nosocomial pathogen spread, combined with fine-grained longitudinal data on human close-proximity interactions (CPIs), we show how detailed knowledge of the structure of human interactions may help design more effective interventions for HAI control. We illustrate this point through an application to control the spread of colonisation by MRSA in a long-term care facility (LTCF).

## Results

### A simulated hospital contact network that realistically mimics the observed contact network

We designed a stochastic individual-based model (IBM) to reproduce the realistic dynamic network of within-hospital between-human interactions. CPI data was collected by equipping all patients and hospital staff in a French LTCF with proximity log-sensors over 84 days (i-Bird study [11,12]). The model was then calibrated to generate simulated contact networks with the same characteristics as the real network provided by the CPI data (see [13] for details). As shown on Figure 1a, the simulated contact network accurately reproduced real average hourly patterns of patient-to-patient, staff-to-staff and staff-to-patient interactions.

**Figure 1:**
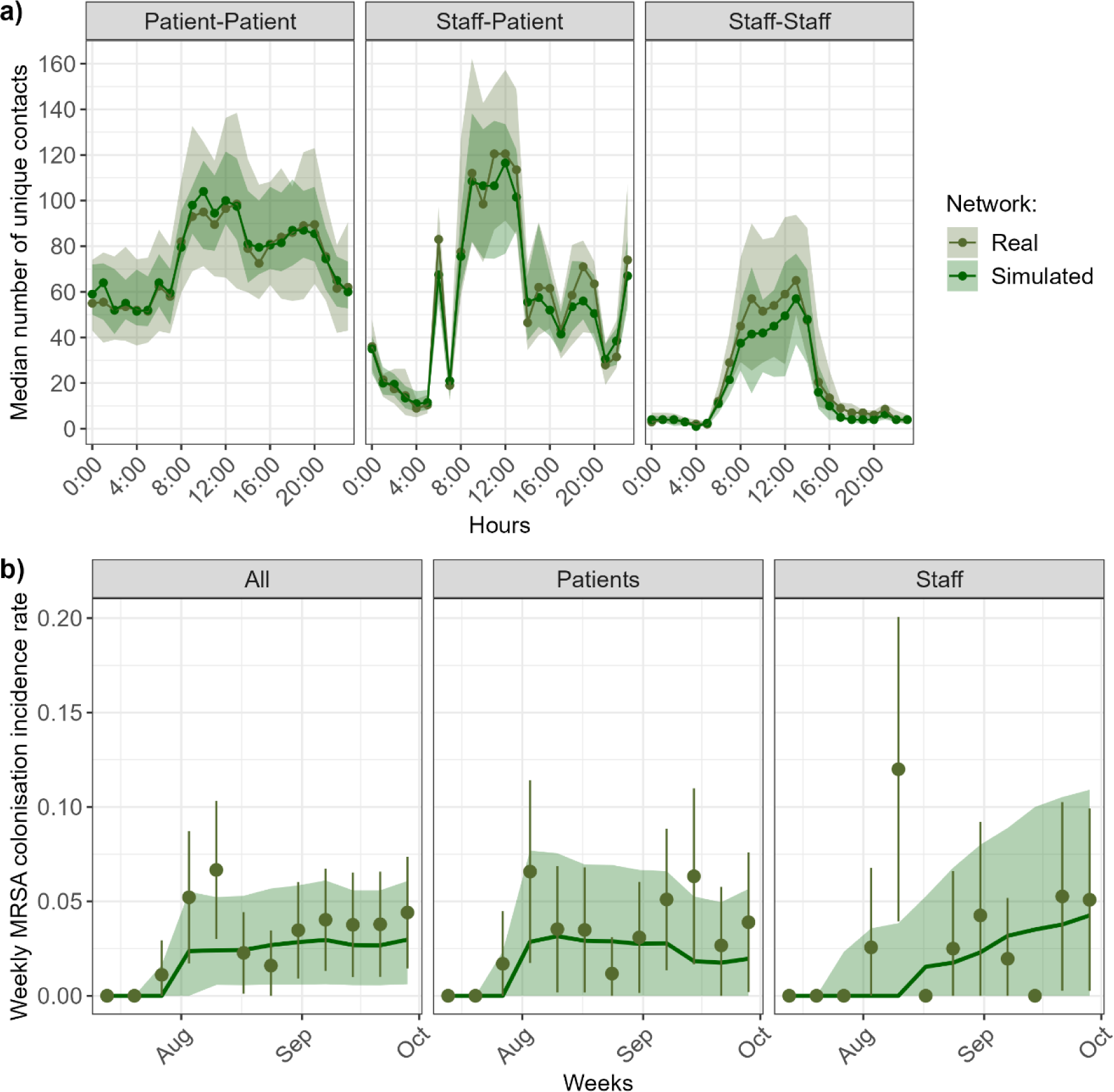
Real and simulated contacts and MRSA incidence. (A) Hourly distribution of number of unique contacts. The lines and points show the median estimates, and the shaded areas show the interquartile ranges. The real values come from the i-Bird study, and the simulated values are shown for 50 simulated contact networks. **(B) MRSA colonisation weekly incidence over 3 months.** Olive points correspond to the observed weekly incidence during the i-Bird study, with lines indicating the margin of error, estimated using the number of individuals swabbed that week. Simulated results are obtained from 15,000 stochastic model simulations (500 simulations of 50 simulated networks). The dark green line shows the median incidence, and the shaded area shows the 95% prediction interval, defined as the interval between the 2.5^th^ and 97.5^th^ percentiles.

### Observed weekly MRSA incidence is well reproduced by simulations of network-based transmission

A Susceptible-Colonised process was implemented into the IBM to reproduce the transmission process of a colonising pathogen, here MRSA, in the LTCF. The model was parameterized to mimic the i-Bird study conditions. An initial 151 patients and 236 hospital staff members were followed up over 84-day simulations. We categorised hospital staffs into 6 groups, (i) healthcare assistants, (ii) nurses, (iii) rehabilitation staff, (iv) physicians, (v) hospital porters and (vi) other. Each day, staff presence and patient admissions and discharges were also simulated using the real data from the i-Bird study. The simulated dynamic contact network described in the previous section was used to mimic between-human interactions and assumed to be the support of MRSA transmission within this LTCF [11,14]. When initializing the model with MRSA carriage of patients and staff as reported by the i-Bird data, the weekly incidence of MRSA colonization predicted by the model reproduced well the observed trends and weekly incidence over the study period (Figure 1b).

### Hospital staff reallocation, especially in healthcare assistants, reduces MRSA spread

To assess the extent to which the dissemination of MRSA can be restricted through an optimized patient-staff allocation, we designed a series of interventions formalized as modifications of the contact network. We assessed the impact of staff reallocation, defined as the attribution of a reduced number of patients to each staff member during the entire investigation period. We maintained the global care needs of patients over the entire period, defined by the number of unique contacts in the data between patients and different staff categories, by ward. A series of scenarios exploring different combinations of staff categories affected by reallocation were implemented and, for each scenario, 30 new contact networks were generated.

Simulating the transmission of MRSA over the different networks, we found that reallocation scenarios targeting different hospital staff categories can help reduce cumulative incidence of MRSA colonisation (Figure 2a for scenarios where 1, 2, or all staff categories were reallocated, Supplementary Figure 1a for all scenarios). Importantly, the benefit of the intervention varied depending on the categories of staff reallocated. When only a single staff category was reallocated, the highest incidence reduction was obtained for healthcare assistant reallocation (median decrease: 10%, 95% confidence interval: 9–11). All scenarios with two categories reallocated involving healthcare assistants prevented between 10–20% of colonisations over the entire simulation period. For comparison, reallocating all staff categories prevented 39% of colonisations (CI: 38–40). Reallocation of either porters or physicians alone barely led to any change in incidence compared to baseline, since these interventions did not substantially change the number of unique staff-patient contacts within the hospital and, therefore, did not substantially affect MRSA spread (Supplementary Figure 2). A pseudo-random contact network in which patients were homogenously distributed among all staff members led to more contacts and a higher incidence as compared to the one generated by the baseline network (36% increase, CI: 35–37), since this increased unique staff-patient contacts within the hospital (Supplementary Figure 2).

**Figure 2:**
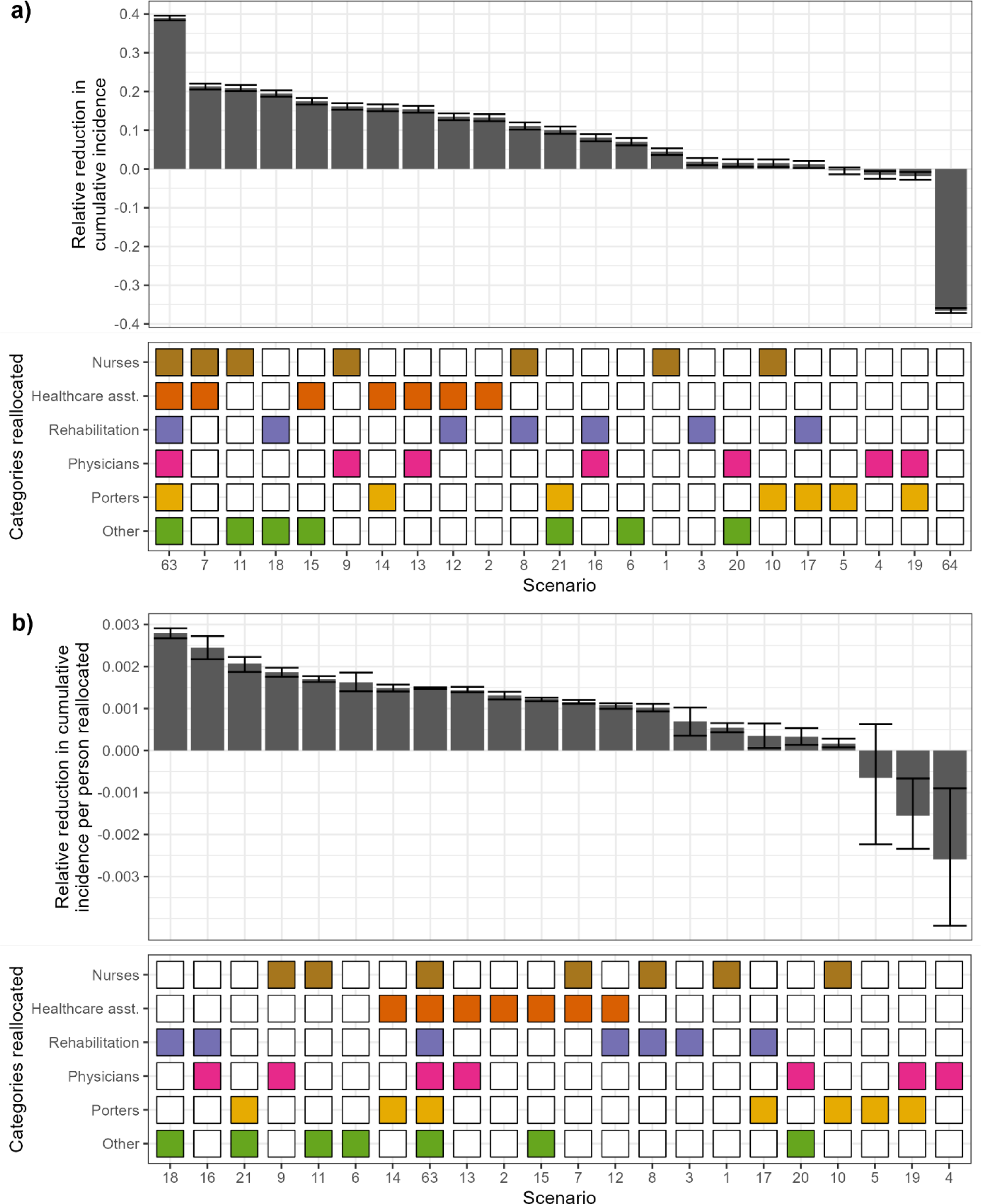
Relative reduction in cumulative incidence of MRSA colonisation for different hospital staff reallocation scenarios, shown per scenario. **(a), or per scenario divided by number of staff reallocated in that scenario (b). Top:** Each bar depicts, for a given scenario, the median relative reduction between 500 model simulations with no intervention, and 500 simulations with staff reallocation, along with the 95% confidence interval. A negative reduction indicates that the intervention led to an increase in cumulative incidence. **Bottom**: In each scenario, staff categories coloured are those reallocated. In scenario 64, the contact network is random. In each plot, the scenarios are ranked from most to least effective.

To see if the variability between scenarios was due to the different number of individuals reallocated in each scenario, we divided the relative incidence reduction for each scenario by the corresponding number of staff reallocated (Figure 2b). This changed the order of the scenarios with the highest benefit, now calculated as relative incidence reduction per reallocated staff. Scenarios where nurses or healthcare assistants were reallocated were lower in the ranking, since they required a large number of staff to be allocated. On the other hand, reallocation of rehabilitation staff and other staff led to the highest overall relative reduction per staff reallocated (2.7 x 10^-3^ %, CI: 2.6 x 10^-3^–2.9 x 10^-3^), even higher than if all staff are reallocated (1.4 x 10^-3^ %, CI: 1.4 x 10^-3^–1.5 x 10^-3^). In any case, we still note heterogeneity in the efficacy of different scenarios, indicating that there are other relevant characteristics which differ between staff categories.

### Reinforced contact precautions of nurses or healthcare assistants are more effective than staff reallocation or vaccination

Next, we investigated the impact of reinforced contact precautions taken by hospital staff (e.g., glove wearing or improved hand hygiene compliance) and vaccination. Contact precautions were simulated as a 2- to 10-fold reduction in both patient-to-hospital staff and hospital staff-to-patient MRSA transmission probabilities during contacts. Vaccination was simulated as a 2- to 10-fold reduction in MRSA acquisition probabilities during contacts between any colonised individual and a non-colonised vaccinated individual. As for the previous analysis, MRSA transmission dynamics were simulated for the different scenarios of reinforced contact precautions and vaccination in the 6 hospital staff categories (Figure 3).

**Figure 3:**
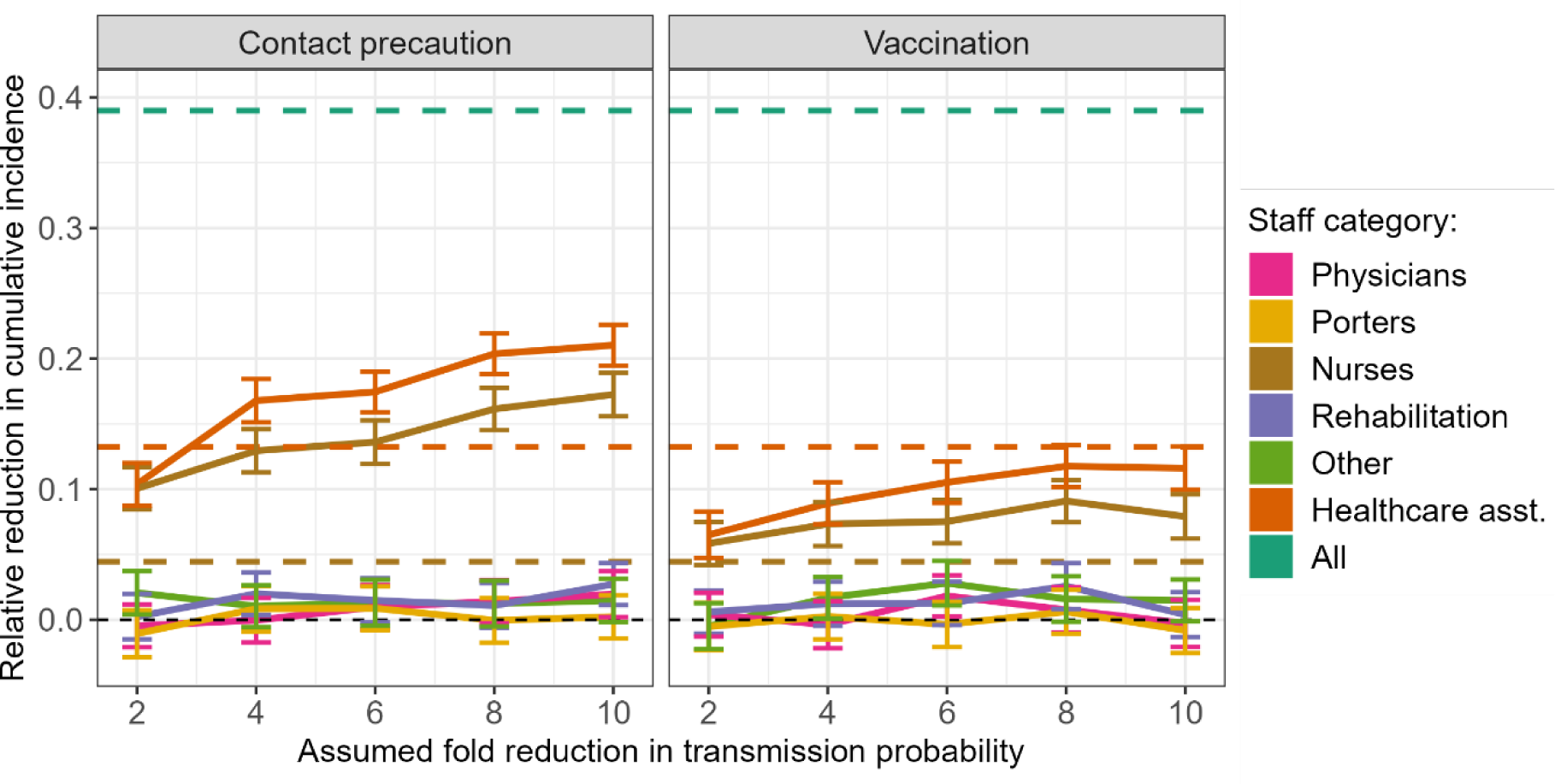
Effect of contact precautions and vaccination targeting different hospital staff categories, compared to staff reallocation. The dashed lines show the median reduction when reallocating all staff (turquoise), healthcare assistants only (orange), or nurses only (brown). All other estimates are shown as median with 95% confidence interval calculated for 500 intervention simulations.

Contact precautions targeting healthcare assistants led to a large reduction in MRSA colonisations, ranging from 10% to 21% as the assumed level of reduction in transmission probabilities increased from 2 to 10-fold (Figure 3). This was closely followed by contact precautions targeting nurses (10-18% reduction). Contact precautions for healthcare assistants or nurses appear to be more effective than reallocation of either of those staff categories alone, as even an assumed 4-fold reduction in transmission probabilities was sufficient to achieve a decrease in incidence slightly higher than reallocation (Figure 3). Whilst vaccination of healthcare assistants or nurses did reduce incidence, the reduction ranged from 6 to 12%, which is approximately equivalent to reallocation (Figure 3).

By opposition, contact precautions or vaccination focused exclusively on either hospital porters, physicians, rehabilitation or other staff appeared ineffective, with percent reductions below 5% irrespective of the assumed transmission probability reduction (Figure 3).

### Heterogeneous distribution of “supercontactors” amongst patients and staff

To understand why intervention effectiveness to reduce the spread of MRSA varied depending on the staff category targeted, we examined the extent to which different individuals were connected in the contact network. We identified individuals substantially more connected than others, and henceforth refer to them as “supercontactors”. We distinguish between two types of supercontactors: (i) individuals with the highest number of daily distinct contacts (henceforth called “frequency-based supercontactors”) and (ii) individuals with the highest overall daily contact duration (henceforth called “duration-based supercontactors”).

We identified the top 60 duration- and frequency-based supercontactors for both patients and staff (i.e. top 20% of individuals). If all individuals had the same probability of being supercontactors, we expect that the distribution of patients/staff categories amongst supercontactors (Figure 4, red and blue) would be aligned with the distribution of those same categories amongst all patients/staff (grey).

**Figure 4:**
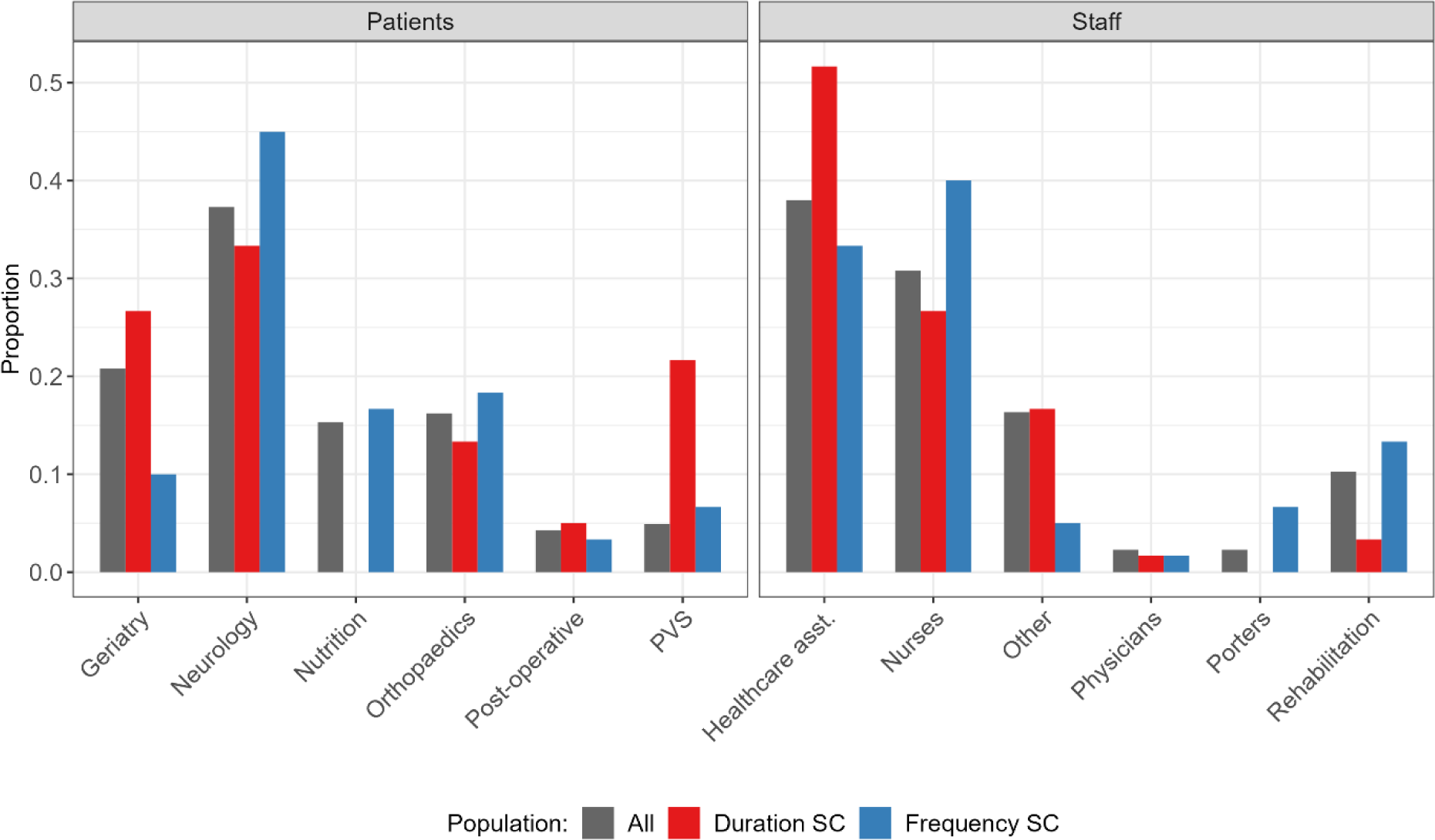
The distribution of supercontactors (SC) amongst hospital patients and staff is not homogeneous. The grey bars show the distribution of categories amongst all patients (left) or staff (right), the red bars show the distribution of duration-based supercontactors, the blue bars show the distribution of frequency-based supercontactors. If supercontactors were homogeneously distributed amongst categories, all the coloured bars would be aligned with the grey bars. Here, only the distribution of the top 60 frequency-based and duration-based supercontactors for patients and staff is shown. PVS: persistent-vegetative state.

Amongst patients, neurology patients are the first category of supercontactors (Figure 4, left; 34% of duration-based, 45% of frequency-based). The observed distribution of patient categories amongst duration-based supercontactors (red) differed significantly from the distribution of those categories amongst all patients (grey; log likelihood ratio test: p value < 0.001). We observed a greater proportion of PVS and geriatric patients amongst duration-based supercontactors than amongst all patients (Figure 4, left). The difference was not statistically significant for frequency-based supercontactors (log likelihood ratio test: p value > 0.2).

Amongst staff, the majority of supercontactors were either healthcare assistants (Figure 4, right; 52% of duration-based, 33% of frequency-based) or nurses (Figure 4, right; 26% of duration-based, 40% of frequency-based). The observed distribution of staff categories amongst supercontactors differed significantly from the distribution of those categories amongst all staff (log likelihood ratio test: duration-based p value < 0.01, frequency-based p value < 0.05). Compared to the distribution amongst all staff, we observed a greater proportion of healthcare assistants amongst duration-based supercontactors, and a greater proportion of nurses, porters and rehabilitation staff amongst frequency-based supercontactors (Figure 4).

There was almost no overlap between the identities of the frequency and duration-based supercontactors. Only three persistent-vegetative state patients, two neurology patients, one nurse and one rehabilitation staff were in both categories.

### Targeting supercontactors is most effective to reduce MRSA spread

We used supercontactors as target for interventions in the hospital. We compared the effect of reinforced contact precautions or vaccination, targeting different combinations of 60 supercontactors (*i.e*. contact-based or duration-based supercontactors among both patients or hospital staff), 60 staff randomly chosen, or 60 patients randomly chosen. Here, we only show the reductions for an assumed 6-fold reduction in transmission probabilities, with other fold reductions shown in Supplementary Figure 3.

Targeting supercontactors within either staff or patients with an intervention was at least as effective to reduce incidence than randomly targeting individuals in the same group with the same intervention (grey, Figure 5). When selecting duration-based supercontactors (red), vaccination targeting patients led to a higher reduction in MRSA colonisations than any intervention targeting hospital staff (Figure 5). Conversely, when selecting frequency-based supercontactors (blue), contact precautions targeting hospital staff gave better results in terms of MRSA colonisations reduction than any intervention targeting patients (Figure 5). Targeting a mix of half frequency- and half duration-based supercontactors (purple) gave intermediary results (Figure 5). Regardless of the type of supercontactors targeted, reinforced contact precautions were more effective than vaccination when targeting staff, whilst vaccination was more effective than contact precautions when targeting patients (Figure 5).

**Figure 5:**
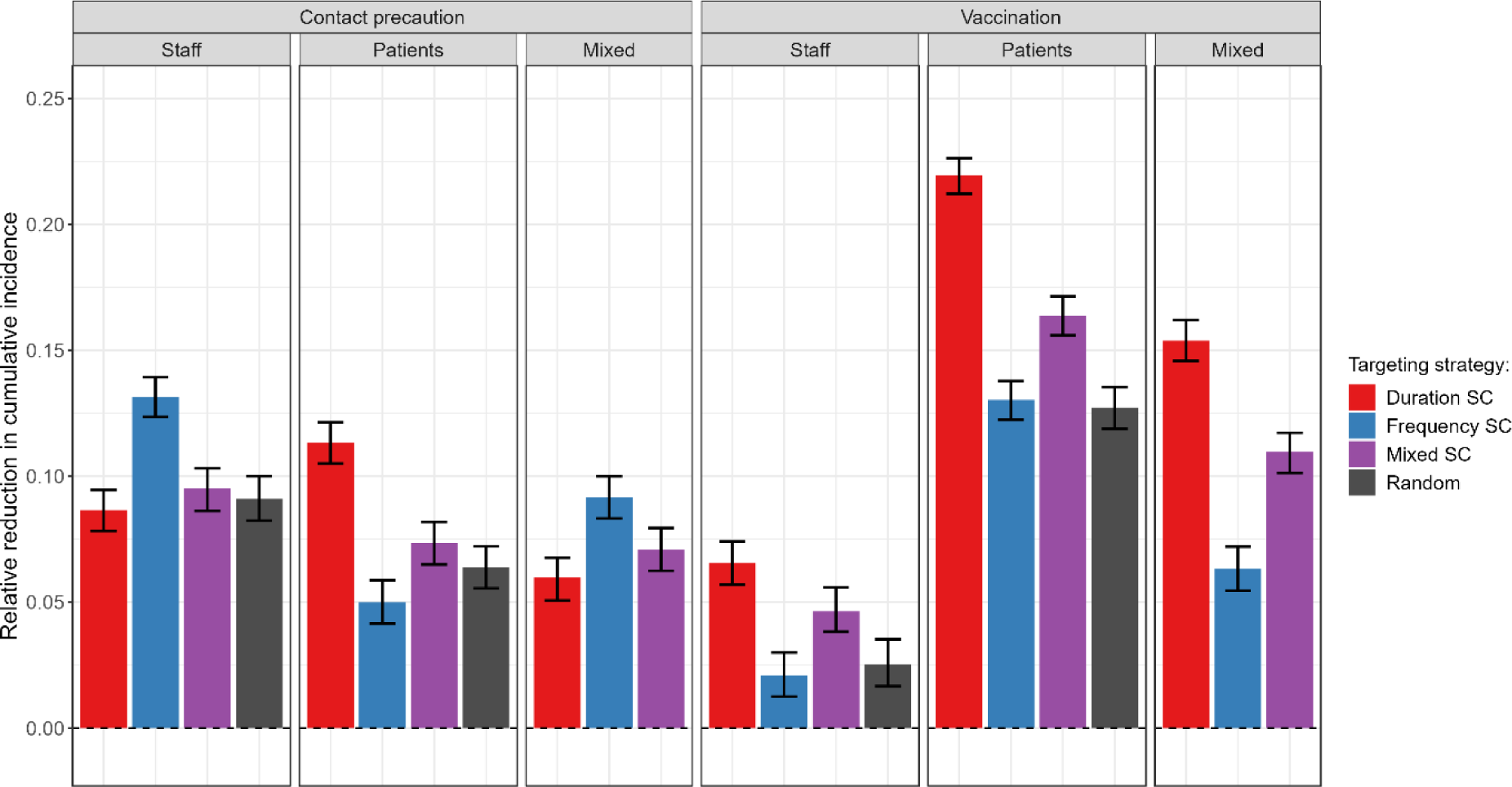
Comparison of contact precautions or vaccination for 60 staff, patients, or a mix of staff and patients, targeting either duration-based supercontactors (SC), frequency-based SC, a mix of duration and frequency-based SC, or random individuals. We assume the interventions lead to a 6-fold reduction in transmission probabilities. For each strategy, the bar indicates the median relative reduction in cumulative incidence, with 95% confidence interval, obtained for 500 simulations.

Overall, vaccination of duration-based patient supercontactors appeared to be the most effective, with up to 21% (CI: 20-22%) of colonisations prevented. These conclusions are maintained when assessing different contact precautions or vaccination efficacies, i.e. assuming 2, 4, 8 or 10-fold reductions in transmission or acquisition probabilities, respectively (Supplementary Figure 3), or when targeting 20 or 100 individuals instead of 60 (Supplementary Figure 4).

### Targeting supercontactors is also an effective strategy for other nosocomial pathogens

Although the epidemiological parameters we used in the previous sections were directly estimated using data on MRSA, our model can be applied to any nosocomial pathogen for which close-proximity interactions are the main vector of transmission. Naturally, the epidemiology of such pathogens would likely vary compared to MRSA, with different transmission rates and carriage/infectiousness durations compared to the values we estimated. To investigate the applicability of our results to other pathogens, we repeated our analysis above, doubling or halving either the transmission rates or the carriage/infectiousness durations. Our qualitative results on the value of targeting supercontactors to improve intervention effectiveness remained valid (Figure 6a-d), even with different baseline incidences due to the parameter changes (Figure 6e). Interestingly, we see that in a few scenarios targeting patients randomly could be slightly more effective than targeting frequency-based patient supercontactors (Figure 6a-d). This is due to the high effectiveness of targeting duration-based patient supercontactors in such instances, combined with the non-overlapping identities of duration- and frequency-based supercontactors. Inevitably, by exclusively targeting frequency-based supercontactors we exclude duration-based supercontactors, while random targeting may still incidentally include these individuals.

**Figure 6:**
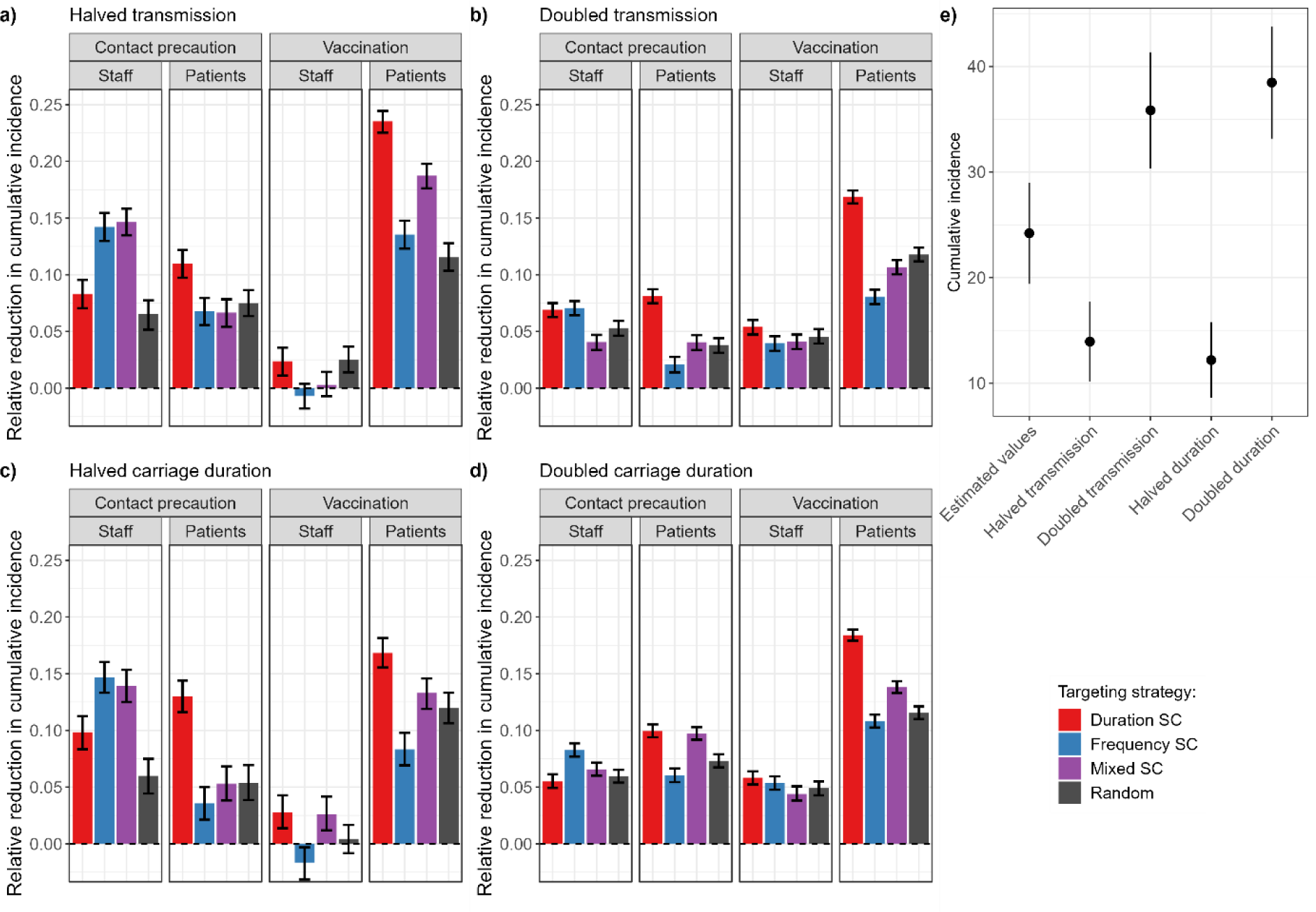
Comparison of contact precautions or vaccination for 60 staff or patients, targeting either duration-based supercontactors (SC), frequency-based SC, a mix of duration and frequency-based SC, or random individuals, and varying either the baseline transmission rate or carriage duration. **a) Halved transmission rate; b) Doubled transmission rate; c) Halved carriage duration; d) Doubled carriage duration.** We assume the interventions lead to a 6-fold reduction in transmission probabilities. For each strategy, the bar indicates the median relative reduction in cumulative incidence, with 95% confidence interval, obtained for 500 simulations. **e) Absolute cumulative incidence without intervention using estimated parameter values, higher/lower transmission, or higher/lower carriage duration.** Points indicate the mean, and lines mean +/- standard deviation, obtained for 500 simulations.

## Discussion

In this study, we present how the dynamic interindividual contact network of a healthcare institution can be analysed to implement efficient interventions aimed at reducing pathogen transmission. We first applied an individual-based model to a French long-term care facility and confirmed that it reproduced well both the recorded network and MRSA dynamics. We then evaluated and compared several network-based control strategies, demonstrating that while hospital staff reallocation can help reduce MRSA transmission overall, staff contact precautions and vaccination could be as or more effective than reallocation. Interestingly, the efficacy varied depending on which staff category was targeted by the intervention. We identified “supercontactors” in the contact network with more or longer contacts and found that these were heterogeneously distributed amongst staff and patient categories. The effectiveness of contact precautions and vaccination was further increased by targeting these supercontactors in the LTCF, compared to randomly targeting individuals. Our conclusions remained valid when varying epidemiological parameters, suggesting that targeting supercontactors is also an effective strategy for other nosocomial pathogens transmitted via close-proximity interactions.

Here we demonstrated that staff reallocation is an efficient strategy to reduce transmission risk. Moreover, reallocation strategies involving healthcare assistants were the most effective. Our simulation results are consistent with previous work on this topic, showing the best staff reallocation strategies were those significantly lowering the degree of the hospital worker-to-patient subgraph [10,15–21]. In a previous study, we examined the potential of different hospital staff categories to spread nosocomial pathogens and to play a role of super-spreader, showing the importance of adherence to contact precautions in “peripatetic” hospital staff. These later were defined as hospital staff members with relatively short contacts, but with many patients, a definition similar to the “frequency-based supercontactors” here [8].

Since transmission was modelled through the contact network, supercontactors can mechanistically play the role of super-spreaders, but also be themselves more at risk of acquiring the bacteria during a contact with a colonised individual. These factors explain why targeting supercontactors for interventions led to a substantial reduction in colonisation incidence. The most appropriate supercontactor type to target (duration-based or frequency-based) surprisingly differed between patients and hospital staff: while targeting frequency-based supercontactors was more relevant for hospital staff, duration-based supercontactors were selected for patients. We also predicted that the most effective intervention to reduce the overall incidence of colonisation was to vaccinate duration-based supercontactors amongst patients with a vaccine, which here we assume protects against acquisition. Interestingly, in staff, vaccination was less effective than reinforced contact precautions. These results may be specific to the type of hospital investigated here. In LTCF, the frequency and duration of patient-patient interactions are much higher than in acute care facilities. Our results highlight the necessity of involving patients in intervention implementation in LTCF.

It is important to note that the hospital followed up during the i-Bird study included neurologic wards hosting patients in persistent vegetative state (PVS). These PVS patients accounted for one fifth of the individuals classified as duration-based supercontactors (Figure 3). While they may be considered similar to sedated and ventilated patients in intensive care units, the presence of this type of patients with particularly long contacts and specific behaviours is not universal across all types of LTCF. To improve the generalisability of our results to other LTCF, we performed an additional analysis in which PVS patients were excluded when identifying supercontactors: this hypothesis did not affect our conclusions (Supplementary Figure 5).

The results presented here should be interpreted in the light of the following limits. Firstly, we only considered here that MRSA transmission occurred through inter-individual contacts among participants, with a risk of transmission saturating after one hour. This assumption was based on previous analysis of the same data, suggesting that the proximity network was the main transmission route for MRSA acquisition in this setting [11]. In this study, whilst participation was high (95% of staff and patients agreed to wear the sensors), it was also estimated that 25% of MRSA acquisitions were not explained by the contact network, and may instead be mediated by other acquisition mechanisms not included in our model, such as environmental contamination, or bacterial evolution within the host leading to the emergence of resistance. Importations of new colonisations, through for example hospital visitors or patient’s permissions outside the hospital were also not included in the model, while they could also have been sources of MRSA acquisition during the i-Bird study. This may explain why model simulations slightly underestimated the incidence point on the 6^th^ week, as illustrated in Figure 1b.

Secondly, we did not account for the infection status of patients in the model. Over the study, several infections occurred in participating patients (eschar, cutaneous infection, gastrostomy, colostomy, tracheotomy, ulcer etc.). When an infection occurs, bacterial load is usually much higher, which could potentially increase the risk of bacterial dissemination in the environment or transmission to contacts. Infections could also impact the dynamic of contacts and of nurse scheduling, as infected patients are bound to have a higher care load, thus requiring more contacts. Interestingly, this higher care load could reclassify infected patients as supercontactors and, as we have shown here, identify them as key targets for interventions to reduce spread. For these reasons, future work taking into consideration infected patients may further improve our ability to implement effective interventions.

Lastly, the epidemiological parameters of the model, which included transmission probability and carriage duration, were directly estimated for MRSA from the admission, schedule, swab and contact data [11,12]. However, these parameters can vary depending on the estimation period (e.g. holidays versus term-time), setting (e.g. long-term versus acute care), population (e.g. older versus younger), and circulating bacterial or viral pathogen in the hospital. For example, the probability of MRSA transmission that we estimated is slightly lower than in other studies (e.g. 0.000023 per 30 seconds of contact on average for hospital staff-to-hospital staff and 0.000789 for hospital staff-to-patient in our study with the real RFID network, compared to a probability between 0.0005 and 0.0050 per 30 seconds of contact in the study by Hornbeck et al [22]). The durations of MRSA colonization that we estimated from the data (31 days for patients, 27 days for hospital staff) are also either shorter or longer than previously reported estimates, but these values can be clone or setting-specific [23,24]. Among other pathogens transmitted by close-proximity interactions, *Klebsiella pneumoniae* has characteristics within the range we explored in our analysis (transmission probability of 0.0005 per 30 seconds of contact, carriage duration of 3 weeks) [25]. SARS-CoV-2 is another example with a similar transmission probability, although the infectious period (equivalent to the carriage duration) is lower (9 days) [26]. As we have shown, our conclusions on the value of interventions strategies targeting supercontactors were not impacted by changes in parameters to reflect the epidemiology of these other pathogens instead of MRSA.

Despite their limitations, mathematical models are powerful tools to inform the efficacy of control strategies in hospital settings [27], when they are based on a good understanding of pathogen transmission routes and heterogeneity in human interactions [28,29]. Over the last decades, different approaches have been used to acquire knowledge on interindividual contacts, such as observational studies, diaries, interviews and more recently wearable sensors [30–38]. While several IBMs of pathogen spread within hospitals [28,39–47] have been developed to assess measures such as hygiene compliance [22] or antiviral prophylaxis impact on influenza [48], few models have actually attempted to directly integrate such rich empiric data. To our knowledge, only two published individual-based models simulated transmission along an RFID-based contact network [11,22], one of which studied MRSA spread [22]. In that work, Hornbeck et al. showed that the number of colonised patients increased when the most connected nurses did not comply with infection control recommendations, which is consistent with our results.

We must consider the feasibility, cost and social acceptability when deciding which control strategies should be implemented. For instance, we suggest that the best strategy would be to implement contact precautions or vaccination focusing on supercontactors, but identifying and targeting supercontactors, in particular among patients, may not be as socially acceptable as broadly targeting hospital staff categories. The benefit of patient vaccination, which we identified as the best strategy in the LTCF, may also be reduced in acute care settings, due to shorter patient lengths of stay and to the likely delay required for immunity to develop following vaccination. In addition, here we chose to simulate the impact of the vaccine as a reduction in the probability of pathogen acquisition, but alternatives could be considered based on recent clinical trials, for example with the vaccine reducing the risk of infection rather than colonisation or reducing the risk of transmission from vaccinated individuals [49]. In any case, achieving a 10-fold reduction in transmission probabilities with these interventions might not actually be feasible, depending on the baseline level of pathogen transmission, which is why we explored a range of reductions as previous studies have done [50]. On the other hand, reallocation requires greater logistical efforts, and may not always be feasible depending on the economic context of the healthcare institution and the care load. Finally, the most effective reallocation strategies may not be the most “cost-effective”. For instance, when considering the relative reduction in incidence per staff reallocated, targeting only rehabilitation staff ranked higher than targeting all staff (Figure 2b).

In conclusion, this work sheds light on the importance of targeting control and prevention measures in an LTCF towards specific hospital staff categories, but also of involving patients in such efforts as they may too play an important role in the transmission network. Patients need to be actors of their own prevention especially when their length of stay is long. More importantly, we underline how monitoring contacts can be helpful to design highly effective control strategies aimed at “supercontactor” individuals.

## Materials and methods

### Data description

Data used here were previously collected during the Individual-Based Investigation of Resistance Dissemination (i-Bird) study [11,12], which took place within a rehabilitation and LTCF from the beginning of July to the end of October 2009. Over this period, each participant (patient or hospital staff) was wearing an RFID sensor that recorded close-proximity interactions (CPIs, at less than 1.5m) every 30 seconds. A dynamic network of proximities is therefore available over 117 days with information on individual ID, ward of affectation, age, gender etc. In addition, dedicated nurses swabbed patients and hospital staff each week to detect MRSA colonization.

The hospital was structured into five wards: (i) three neurological wards, (ii) one nutritional care ward and (iii) one geriatric ward. In addition to neurologic, geriatric and nutritional care patients, the hospital hosted a few persistent vegetative state (PVS), post-operative and orthopaedic patients. Most patients had long hospitalization durations (median: 7 weeks). In addition to “classic” staff categories such as nurses, physician, rehabilitation staff, patients could interact with other staff members, such as hairdressers.

Overall, a total of 327 patients and 263 hospital staff had recorded contacts during the investigation period. This study is described in more detail in [11,12].

### Model description

We developed a stochastic Susceptible-Colonized-Susceptible individual-based model that simulates the dynamic transmission of a pathogen within a hospital over a network incorporating data on the detailed structure of CPIs. Individuals could either be patients or hospital staff members. Hospital staff were divided into six categories: healthcare assistants, nurses (including nurses, head nurses, and students), rehabilitation staff (occupational therapists, physiotherapists, and other rehabilitation staff), physicians, hospital porters, and other staff (animation, logistic, administration, and hospital service agents). The model accounts for admissions and discharges from the hospital and inter-individual contacts. Once admitted, a patient remains in the hospital until discharged, whereas hospital staff can be present or absent according to their daily schedule. A probability per time unit for hospital staff presence simulates this schedule.

#### Transmission process

Every individual can either be colonized or non-colonized (susceptible) by the pathogen (here, MRSA). At each contact between a susceptible and a colonized individual, the pathogen can be transmitted from the colonized to the susceptible individual with a given probability. This transmission probability is computed as the product of the between-individual contact duration and the pathogen-specific transmission probability, assuming that risks saturate after 1 hour. The model accounts for four different transmission probabilities depending on the status of the individuals involved: patient-to-patient, patient-to-staff, staff-to-patient and staff-to-staff (see Supplementary Text 1). A colonized individual can naturally recover to the susceptible state after a colonization duration randomly drawn from a lognormal distribution. Such individuals may subsequently be recolonised (no immunity is assumed). We also assume that no active decolonisation measures are implemented.

Individuals are assumed to be screened for colonization with a probability estimated from the data that depends on weekdays.

#### Model parameterization

The model was parameterized using i-Bird data. Simulations ran over 84 days, with an initial 151 patients and 236 hospital staff members present, to reflect the duration and conditions of the data collection. Values for model parameters were also computed from the observed data on MRSA colonization among the patients and hospital staff. A summary list of model parameters is provided in Table S2. Detailed information on parameter value calculations is provided in Supplementary Text 1.

### Building synthetic contacts

We built an algorithm to generate both realistic full and reported stochastic dynamic networks of interindividual interactions in the hospital using parameters estimated from the observed data. Details of parameters computations and CPI generation algorithm are provided in reference [13].

### Implementing control strategies

We evaluated three distinct contact-based control strategies: staff reallocation, contact precautions, and vaccination.

*Reallocation* was simulated as a modification of the contact network, in which patients were allocated to a given staff member of each category for their entire length of stay. We then generated contacts using the algorithm we previously described [13], choosing in priority the staff member allocated to that patient (or vice-versa) when a corresponding contact occurred. For example, if we allocate patient *p1* to nurse *n1*, then nurse *n1* will systematically be chosen in priority whenever the algorithm attempts to create a contact between *p1* and a nurse. We assumed that reallocation did not influence CPI rates.

*Contact precautions* were simulated by reducing instantaneous patient-to-hospital staff and hospital staff-to-patient transmissions probabilities 2-, 4-, 6-, 8- or 10-fold, irrespective of CPI rates. Three specific scenarios were investigated: (i) contact precautions for all members of different staff categories, (ii) contact precautions for 60 randomly selected staff members amongst nurses or all staff, and (iii) contact precautions for 60 individuals with the highest rates of contacts, called “supercontactors”. Two definitions of supercontactors were assessed: (i) based on the number of contacts (henceforth called “frequency-based supercontactors”) and (ii) based on the duration of contact (henceforth called “duration-based supercontactors”). Frequency-based supercontactors were defined as the patients or hospital staff members who had the highest mean number of daily contacts with distinct individuals. Duration-based supercontactors were defined as the patients or hospital staff members who had the highest mean daily cumulative duration spent in contact with other individuals. Several strategies were explored regarding the type (patients and/or staff members) of selected supercontactors on whom to focus reinforced contact precautions.

*Vaccination* was simulated by reducing acquisition probabilities for vaccinated individuals by 2-, 4-, 6-, 8- or 10-fold, irrespective of CPI rates. This effectively corresponds to an unvaccinated-to-vaccinated transmission probability reduction, regardless of the categories of individuals in contact (staff or patient). For example, a 6-fold reduction would translate into a vaccine efficacy of 1 - 1/6 = 83% to reduce the risk of acquisition. We examined the same scenarios as for contact precautions. We assume that the vaccine has been administered with sufficient time before the simulation, and therefore do not consider a delay before reaching maximum vaccine efficacy. We also do not account for potentially waning immunity due to the relatively short time period of our simulation.

For all interventions and scenarios, the relative reduction in the cumulative incidence of MRSA colonisation over the entire simulation period was used as an indicator of intervention efficacy. This was calculated by simulating each scenario (including baseline) 500 times and comparing each simulation result with 10 randomly chosen simulations of the baseline scenario, leading to a total of 5000 comparison points per scenario. We used a Wilcoxon test to check if the median relative reduction in cumulative incidence was significantly different from 0.

We used the model to simulate the impact of these control strategies for other pathogens than MRSA. To represent the varying epidemiological characteristics of these pathogens, we either doubled or halved the values for the transmission rate or carriage duration (i.e. infectious period) compared to the values we estimated from the data.

## Data Availability

Model results and analysis scripts are available online at https://github.com/qleclerc/ctcmodeler

## Acknowledgements and funding sources

This project has received funding from the Innovative Medicines Initiative 2 Joint Undertaking under grant agreement No 101034420 (PrIMAVeRa). This Joint Undertaking receives support from the European Union’s Horizon 2020 research and innovation programme and EFPIA.

This communication reflects the author’s view and that neither IMI nor the European Union, EFPIA, or any Associated Partners are responsible for any use that may be made of the information contained therein.

## Notes

### Competing Interest Statement

The authors have declared no competing interest.

