## Supplementary Material for "Using contact network dynamics to implement efficient interventions against pathogen spread in hospital settings"

**Classification:** Biological Sciences, Biophysics and Computational Biology

**Keywords:** long-term care facility, contact network, individual-based model, nosocomial pathogens, interventions

### SUPPLEMENTARY INFORMATION

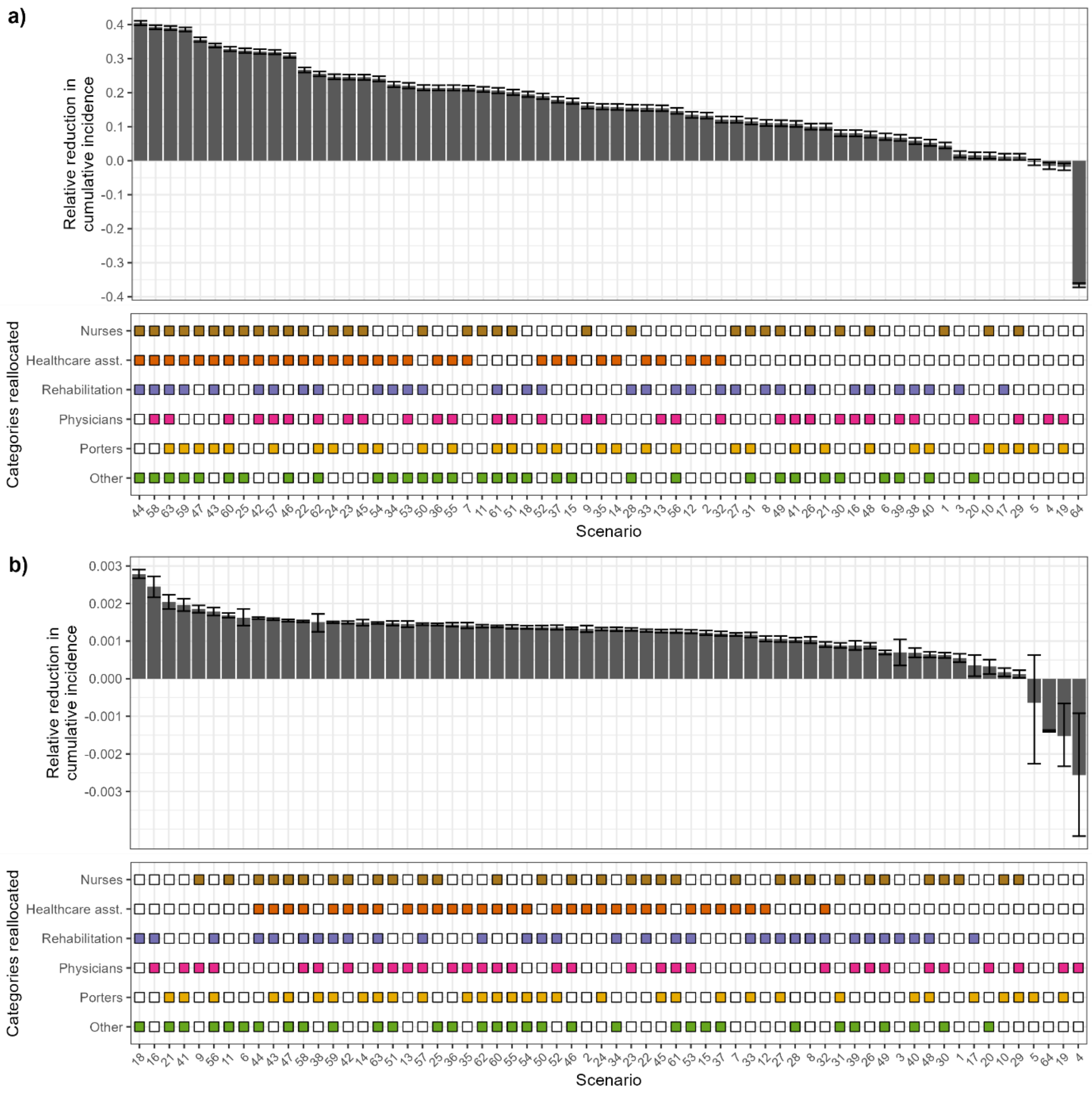


**Supplementary Figure 1 : Relative reduction in cumulative incidence of MRSA colonisation for different hospital staff reallocation scenarios, shown per scenario (a), or per scenario divided by number of staff reallocated in that scenario (b). Top:** Each bar depicts, for a given scenario, the median relative reduction between 500 model simulations with no intervention, and 500 simulations with staff reallocation, along with the 95% confidence interval. A negative reduction indicates that the intervention led to an increase in cumulative incidence. **Bottom**: In each scenario, staff categories coloured are those reallocated. In scenario 64, the contact network is random. In each plot, the scenarios are ranked from most to least effective.

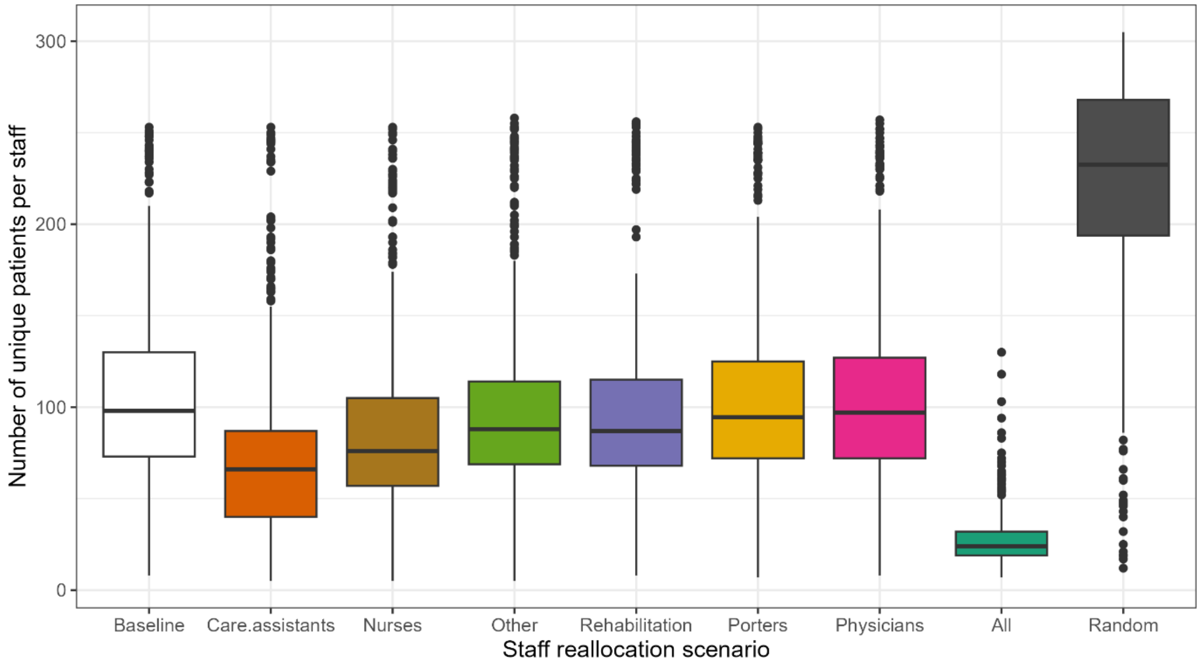


**Supplementary Figure 2 : Number of unique patients in contact with each staff member in the baseline network, compared to reallocation scenarios involving either one staff category at a time, all staff, or a random allocation.**

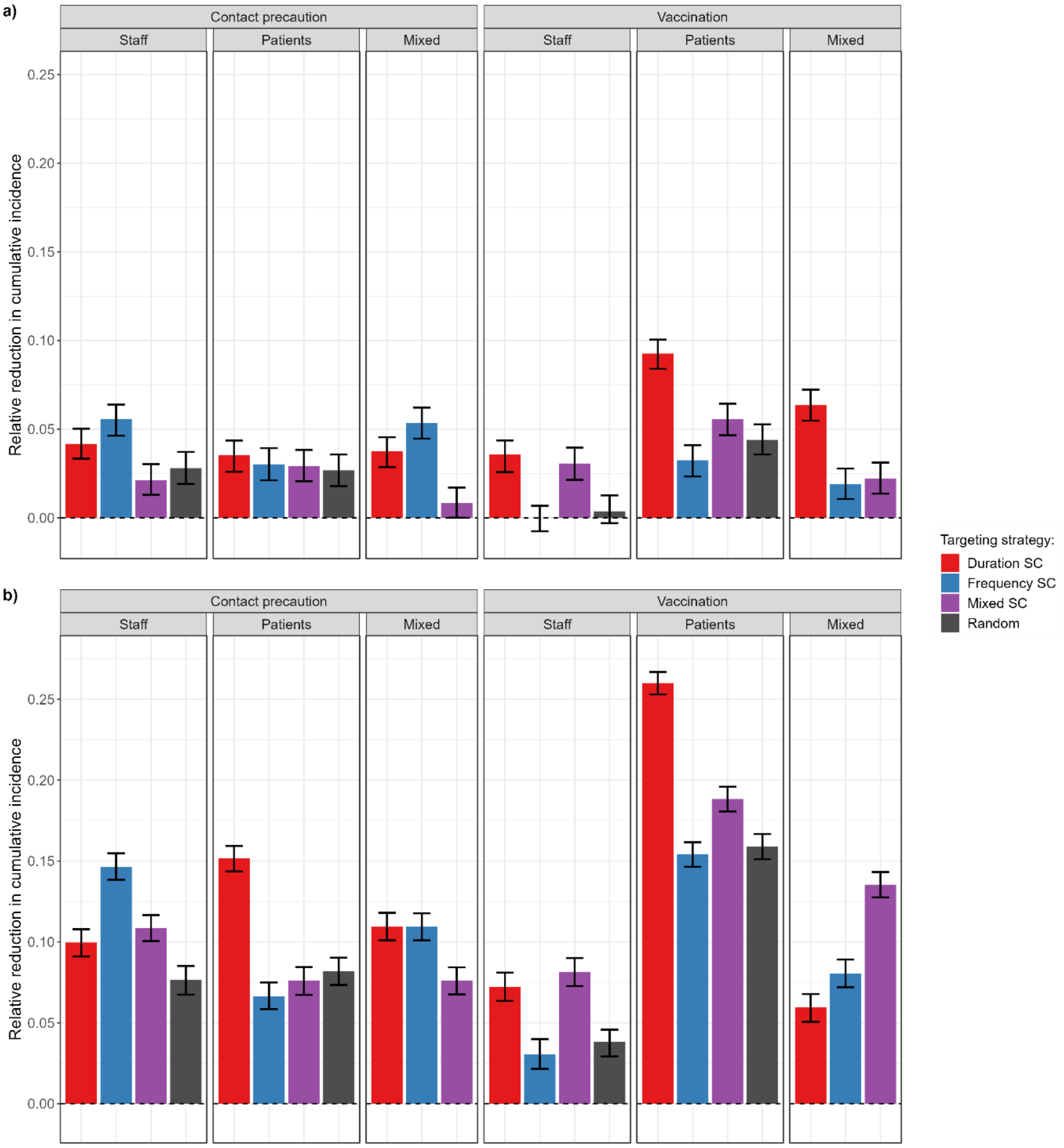


**Supplementary Figure 3: Comparison of contact precautions or vaccination targeting 60 individuals, either selected randomly amongst staff or patients, or different groups of supercontactors, assuming a fold-reduction in transmission probability of a) 2 or b) 10.** For each strategy, the bar indicates the median relative reduction in cumulative incidence, with 95% confidence interval, obtained for 500 simulations.

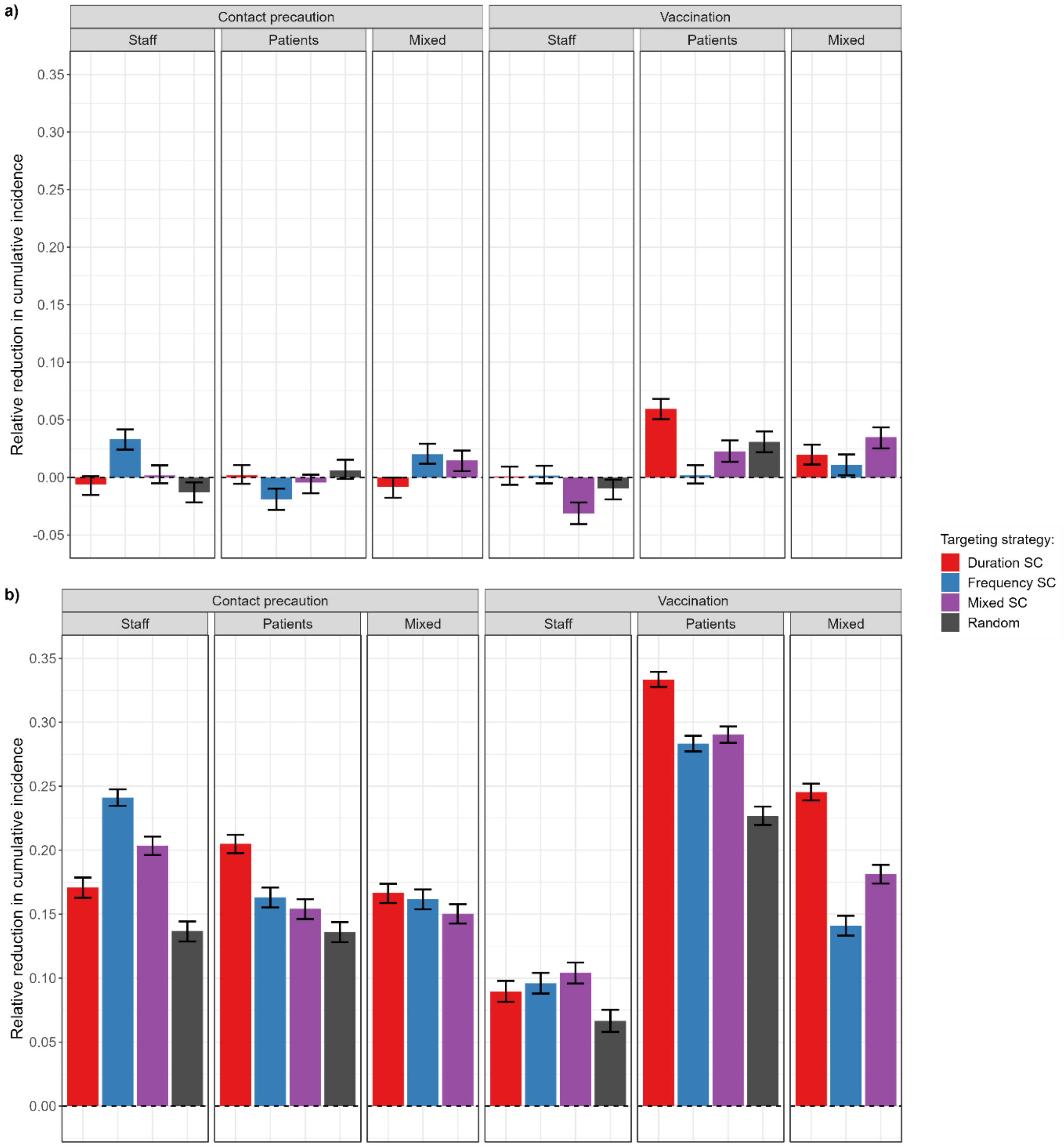


**Supplementary Figure 4: Comparison of contact precautions or vaccination targeting a) 20 or b) 100 individuals, either selected randomly amongst staff or patients, or different groups of supercontactors, assuming a 6 fold-reduction in transmission probabilities.** For each strategy, the bar indicates the median relative reduction in cumulative incidence, with 95% confidence interval, obtained for 500 simulations.

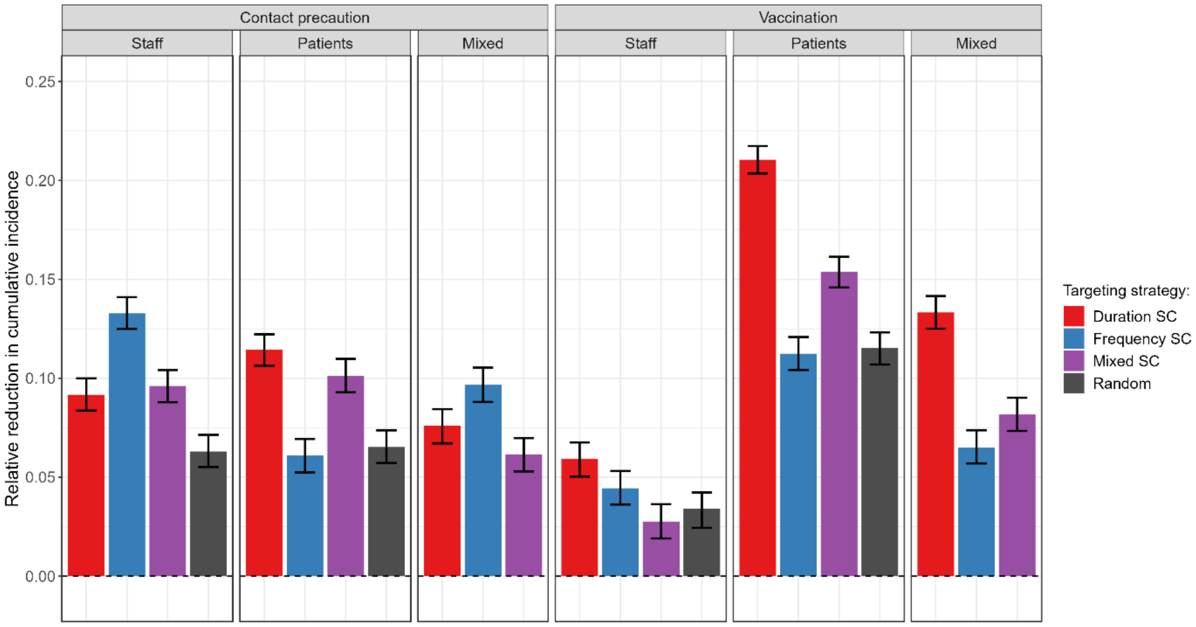


**Supplementary Figure 5: Comparison of contact precautions or vaccination targeting 60 individuals, either selected randomly amongst staff or patients, or different groups of supercontactors excluding persistent-vegetative state patients, assuming a 6-fold reduction in transmission probabilities.** For each strategy, the bar indicates the median relative reduction in cumulative incidence, with 95% confidence interval, obtained for 500 simulations.

### Text S1 – Detailed CTCmodeler description

CTCmodeler is composed of an individual-based model (IBM) previously described [26,51], and a module to compute model parameters.

#### IBM description

The IBM module simulates the nosocomial transmission of a pathogen through an interindividual contact network. Three time schedules are used (i) time steps, (ii) days and (ii) weeks.

(i) The model runs using discrete 30-second time-steps. At each time step, the model simulates contacts and transmission events between individuals according to defined transmission probabilities between the groups these individuals belong to (patients or hospital staff). When an individual newly acquires the pathogen, their status changes from “susceptible” to “colonized”. A colonized individual will stay colonized for a duration sampled from a lognormal distribution at the time of the acquisition, after which their status reverts from “colonized” to “susceptible”.

(ii) Admissions and discharges operate every day. If an admission occurs, the probability that the new individual is admitted with a “colonized” status depends on the defined colonization frequency at admission among patients. Swabbing, that will determine the observation process of colonization status, also operates daily, with a test for each individual to assess whether they are swabbed on that day.

(iii) Every week, the model selects the number of individuals that will be swabbed, and the corresponding swabbing days, according to a normal distribution informed by the average number of swabs taken for different staff or patient categories per day during the i-Bird study. The individuals are then chosen randomly amongst all those belonging to the corresponding category.

#### Outcomes of the model

To take into account a reporting process, the incidences and prevalence are those observed, and hence only include colonized individuals that have been swabbed. Sensitivity of the swab test is assumed to be perfect. The incidence is the number of weekly new acquisitions divided by the number of susceptible individuals the previous week.

#### Parameters estimation from data

Epidemiological parameters of the IBM are computed from the analysis of weekly longitudinal swab data obtained from the LTCF. The following parameters are estimated:

##### *Colonization duration*

The mean and variance of the colonisation duration were computed separately for patients and hospital staff, using data from all the colonisation events recorded in the i-Bird data. The colonisation duration $D_i$ of one colonisation episode *i* was estimated as:

$$D_i = \frac{D_{i,pos} + D_{i,neg}}{2} \quad (1)$$

Where $D_{i,pos}$ is the number of days between the first and the last positive swabs and $D_{i,neg}$ is the number of days between the last and the first negative swabs, respectively before and after those positive swabs.

##### *Colonization probability at admission*

The colonisation probability at admission was estimated for patients and hospital staff separately, by dividing the number of individuals with a positive swab within 48h of admission by the total number of individuals swabbed within 48h of admission.

##### *Distribution of swabs across weekdays*

The average number of swabs for each weekday *j* (i.e. Monday, Tuesday, Wednesday, Thursday, Friday) for each group of individuals *g* (i.e. patient or staff) are computed as:

$$P_{g,j} = \frac{1}{J_j} \sum_{w=1}^{W} R_{g,j}^{w} \quad (2)$$

Where $J_j$ is the total number of specific days *j* (e.g. number of Mondays) over the investigation period, $R_{g,j}^{w}$ is the total number of individuals of group *g* swabbed on a day *j* on week *w*, and

$W$ is the total number of weeks. We assumed that there were no swabs taken on weekends, as observed during the i-Bird study.

***Admission rate***

The admission rates were estimated separately for each group (patient or staff), category (patient reason for hospitalisation or staff category) and ward, by dividing the total number of individuals of each group-category-ward combination admitted to the hospital by the length of the investigation period (84 days).

***Length of stay***

The mean and variance of the length of stay $Md_{g,c,s}$ for individuals of group $g$ (i.e. staff or patient), category $c$ (staff category or patient reason for hospitalization) and ward $s$ are computed as:

$$E(Md_{g,c,s}) = \frac{1}{I_{g,c,s}} \sum_{i=1}^{I_{g,c,s}} \frac{1}{S_i} \sum_{j=1}^{S_i} D_j^i \quad (3)$$

$$Var(Md_{g,c,s}) = \frac{1}{I_{g,c,s}-1} \sum_{i=1}^{I_{g,c,s}} \left[ \frac{1}{S_i} \sum_{j=1}^{S_i} D_j^i - E(Md_{g,c,s}) \right]^2$$

Where $I_{g,c,s}$ is the total number of unique individuals of group $g$, category $c$ and ward $s$ across the study period, $Si$ is the total number of hospital stays for the individual $i$ and $D_j^i$ is the duration of hospital stay $j$ (in days).

***Staff presence rate***

Presence rate $TP_{h,c,s}$ of staff category $c$ at hour $h$ in ward $s$ is computed as:

$$TP_{h,c,s} = \frac{\sum_{i=1}^{N^{cs}} \sum_{k=1}^{N^h} I_{i,k}}{N^h} \quad (4)$$

Where $I_{cr,i}$ is equal to 1 if the staff member $i$ from category $c$ allocated to ward $s$ is present inside the hospital at instance $k$ of the hour $h$ and 0 otherwise. $N^{cs}$ is the total number of staff of category $c$ allocated to ward $s$. $N^h$ is the number of instances of the hour $h$ included in the study period (i.e. the number of days of the study period that contain the hour $h$).

***Transmission probabilities***

Transmission probabilities between two groups *g1* and *g2* (patients or staff) were estimated based on i-Bird contact data and swabs results as follows:

$$T_{g1\rightarrow g2} = \frac{1}{(N^w - n)} \sum_{w=n+1}^{N^w} \frac{\sum_{i=1}^{C^{g1}_{w-n\rightarrow w-1}} \sum_{j=1}^{S^{g2}_{w-n\rightarrow w-1}} I_{i,j,w-n\rightarrow w-1} \times A_{w,j}}{\sum_{l=w-n}^{w-1} D_{C^{g1}\rightarrow S^{g2},l}} \qquad (5)$$

Using n=2 allows us to account for imperfect sensitivity of the swab tests, while n=1 assumes perfect sensitivity. We defined acquisition when one positive swab followed two previous negative swabs. $N^w$ is the total number of weeks. For each week *w*, $C^{g1}_{w-n\rightarrow w-1}$ is the number of *g1* individuals with a positive colonisation status between *w-n* and *w-1* and $S^{g2}_{w-n\rightarrow w-1}$ is the number of *g2* individuals with a negative colonisation between *w-n* and *w-1*. $I_{i,j,w-n\rightarrow w-1}$ is equal 1 if the individual *i* (colonized) and the individual *j* (susceptible) during the period *w-n* to *w-1* were in contact, and 0 otherwise. $A_{w,j}$ is equal to 1 if *j* had an acquisition during week *w*, and 0 otherwise. $Dc_{t1\rightarrow t2,l}$ is the cumulative contact duration between colonized *g1* individuals and susceptible *g2* individuals during week *l*. Only the first acquisition event of an individual was considered.

The resulting parameter values are listed in Supplementary Table S2.

**Supplementary Table S2. List of model parameters used for the agent-based model**

| **Hospital characteristics** | |
|---|---|
| Initial number of patients | 151 |
| Initial number of hospital staff | 236 |
| Number of swabs according to weekdays for patients (range of mean (range of variance)) | 4.45-61.36 (6.67-229.45) |
| Number of swabs according to weekdays for hospital staff (range of mean (range of variance)) | 7.36-24.18 (4.56-102.56) |
| **Pathogen characteristics** | |
| Colonization frequency at admission among patients | 32% |
| Colonization frequency at start among hospital staff | 70% |
| Duration of carriage among patients (mean (variance)) in days | 31.75 (764.36) |

| | |
|---|---|
| Duration of carriage among hospital staff (mean (variance)) in days | 27.10 (557.07) |
| Patient to patient transmission probability per 30 seconds of contact | $2.3 \times 10^{-5}$ |
| Patient to staff transmission probability per 30 seconds of contact | $1.19 \times 10^{-4}$ |
| Staff to patient transmission probability per 30 seconds of contact | $7.89 \times 10^{-4}$ |
| Staff to staff transmission probability per 30 seconds of contact | $1.66 \times 10^{-4}$ |

#### Simulations

For each scenario, 500 independent stochastic simulations were performed. The model was coded in C++ with the repast HPC 2.3.0 library. All simulations were performed on the Maestro cluster hosted by the Institut Pasteur.
